# The transition zone in Hirschsprung’s bowel contains abnormal hybrid ganglia with characteristics of extrinsic nerves

**DOI:** 10.1101/2022.06.15.22276240

**Authors:** Megan Smith, Sumita Chhabra, Rajeev Shukla, Simon Kenny, Sarah Almond, David Edgar, Bettina Wilm

## Abstract

The aganglionic bowel in short segment Hirschsprung’s disease is characterised both by the absence of enteric ganglia and the presence of extrinsic thickened nerve bundles (TNBs). The relationship between the TNBs and the loss of enteric ganglia is unknown. Previous studies have described decreasing numbers of ganglia with increasing density of TNBs within the transition zone (TZ) between ganglionic and aganglionic gut, and there is some evidence of spatial contact between them in this region. To determine the cellular interactions involved, we have analysed the expression of perineurial markers of TNBs and enteric ganglionic markers for both neural cells and their ensheathing telocytes across four cranio-caudal segments consisting of most proximal ganglionic to most distal aganglionic from pull-through resected colon. We show that in the TZ, enteric ganglia are abnormal, being surrounded by perineurium cells characteristic of TNBs. Furthermore, short processes of ganglionic neurons extend caudally towards the aganglionic region, where telocytes in the TNB are located between the perineurium and nerve fibres into which they project telopodes. Thus, enteric ganglia within the TZ have abnormal structural characteristics, the cellular relationships of which are shared by the TNBs. These findings will help towards elucidation of the cellular mechanisms involved in the aetiology of Hirschsprung’s disease.

## Introduction

Hirschsprung’s disease (HSCR) is a gut motility disorder which occurs in 1 in 5000 births and is four-fold more prevalent in males than females [1-3]. It is characterised by the absence of enteric ganglia due to the failure of the enteric nervous system (ENS) progenitor cells to entirely colonise and/or differentiate within the bowel during prenatal development [1]. Short segment HSCR (SS-HSCR) is the most common presentation of HSCR, with 80% of HSCR cases having aganglionosis restricted to the rectosigmoid colon. Less frequently, greater lengths of the bowel can be affected, with up to 10% of cases being total colonic HSCR and 1% being total intestinal HSCR [1,3].

The aganglionic region of SS-HSCR bowel contains extrinsic, abnormally thickened nerve bundles (TNBs) consisting of axons from pelvic ganglia and glial cells [4]. While in HSCR the TNB typically have diameters larger than 40μm [5], nerves with diameters up to 150μm have been described [6,7]. However, it is unknown why these TNBs develop in SS-HSCR [8,9], and specifically whether the lack of ENS in the distal bowel of SS-HSCR is a cause, a consequence, or is unrelated to the presence of TNBs.

In the healthy bowel, p75 is expressed by non-neuronal cells derived from the neural crest. Glucose transporter 1 (GLUT1) is expressed in perineurial cells which as perineurium ensheath the endoneurium of extrinsic nerves [10]. By contrast, GLUT1 is not expressed by cells of the ENS in the healthy bowel [6,10,11], and only very small extrinsic nerves with a GLUT1-positive perineurium are found in the most distal rectum of the healthy gut [12]. The TNBs observed in SS-HSCR have been shown to be surrounded by a GLUT1-positive perineurium, the cells of which also express the low affinity nerve growth factor receptor (p75) [6,13,14]. In TNBs, endoneurial cells also express p75 [15], but not GLUT1.

Enteric submucosal and myenteric ganglia in healthy intestine do not have a perineurium, but instead are ensheathed by a layer of telocytes, interstitial cells which form a cellular network [16], projecting telopodes into the ganglia [17]. The key marker expressed by telocytes bounding enteric ganglia is CD34, a transmembrane phosphoglycoprotein also expressed in haematopoietic stem cells; importantly, telocytes do not express GLUT1 or p75 [18]. The role of telocytes in the gut is not fully understood, but it has been proposed that they provide mechanical stability, regulate extracellular matrix organisation [16,19] and may be mesenchymal precursors of interstitial cells of Cajal (ICCs) during cell turnover [17,20]. Interestingly, extrinsic nerves are surrounded by telocytes in the endo- and perineurium [21]. Despite the potentially important functions of telocytes in the cellular organisation of the ENS, their localization in the aganglionic region of HSCR has not been characterised.

The region between the normal ganglionic and the aganglionic regions of the bowel in SS-HSCR is characterised cranio-caudally by a gradual loss of ganglia, and conversely by increasing numbers of TNBs [10]. Within this region there is a segment called the transition zone (TZ) where both TNBs and enteric ganglia co-exist [10]. The extent of the TZ can vary between children with SS-HSCR with an inverse relationship between extent of aganglionosis and the length of the TZ, whereby in patients with shorter aganglionic region the TZ has a longer length [10,22]. Additionally, smaller GLUT1-positive nerve bundles (with a diameter smaller than 40μm) are found throughout the TZ [10]. Taken together, these observations prompt the question whether there may be direct cellular contacts and interactions between ENS ganglia and extrinsic nerves within the TZ. The characterization of these interactions may help in the elucidation of the relationship between ENS and TNBs relevant to the pathology of HSCR.

The histopathological characteristics applied in clinical practice to distinguish between the TZ and the normally innervated ganglionic region [23], include partial circumferential aganglionosis, hypertrophic submucosal nerves and myenteric hypoganglionosis [10], as determined by GLUT1, calretinin, haemotoxylin and eosin (H&E), and acetylcholinesterase staining. In HSCR bowel, the loss of ganglia cranio-caudally is accompanied by a loss of the expression of the ENS-specific neuronal marker calretinin [24] and of the pan-neuronal marker Hu [10]. However, a detailed analysis of the co-existence of the ENS ganglia and TNBs within the TZ is still missing. Further, it is unclear how the loss of enteric ganglia and an increase in TNB affects the telocytes and perineurial cells. To elucidate interactions between the intrinsic and extrinsic innervation within the TZ we therefore performed immunostaining for key markers at four levels in the pull-through resected colon of children with SS-HSCR. Our analysis details the morphological and cellular relationships between enteric ganglia and the appearance of TNBs along the resected tissue. We also describe the distribution of telocytes in SS-HSCR, from ganglionic bowel through the TZ and in the TNBs of the aganglionic region.

## Materials and Methods

### Patient material

This study was carried out under the ethical approval obtained from the UK North West 3 Research Ethics Committee (Ref:10/H1002/77). Human colon tissue was collected from 4 children with short-segment Hirschsprung’s disease (2 male, 2 female) undergoing a pull-through procedure and were aged between 1-5 months at time of surgery. Of the 4 patients, 3 children underwent primary pull-through while 1 child had a stoma placed before undergoing pull-through surgery. The pull-through resected bowel was divided into 4 segments based on the appearance and morphology of the rectosigmoid pull-through resected tissue as well as the histopathology results of intraoperative biopsies. Full thickness samples were taken from segments identified as I-IV, starting from the ganglionic region and through to the aganglionic region of the rectosigmoid pull-through resected specimen (Supplementary Figure 1). Segment I was identified as a region in which there were ganglion cells present on H&E staining of frozen sections of seromuscular biopsies obtained intra-operatively. Segment IV was identified as the most distal aganglionic region and selected based on macroscopic appearance and narrowed morphology when compared to the more proximal colon, in addition to previous immunohistochemical rectal suction biopsy results. Segments II and III were collected by dividing the tissue between segments I and IV in half, with the most proximal region being segment II and more distal segment III.

### Tissue processing and immunofluorescence

The full thickness tissue specimens were fixed in 4% (w/v) paraformaldehyde for 4 hours before being placed in 30% (w/v) sucrose for cryoprotection and embedded in Shandon Cryomatrix. The specimens were then cryosectioned at 7μm and stored at -20°C.

The following primary antibodies were used for immunofluorescence on serial sections (Supplementary Table 1): Hu, calretinin, GLUT1 and CD34. All antibodies were co-stained with p75. Secondary antibodies used were Alexafluor Goat anti-Mouse IgG1 594 (p75) and Alexafluor Goat anti-Rabbit 488 (all other markers) and counterstained with mounting medium with DAPI (Abcam). Images were taken on a Leica DM2500 upright microscope x40 objective with DFC350FX camera Leica Application Suite software (LAS) Version 4.2, or Zeiss LSM 800 Airyscan laser scanning confocal with x63 oil objective with integrated camera using Zeiss Zen software. All images were then merged and assembled in FIJI (ImageJ).

## Results

### p75 and GLUT1 co-expression in cells surrounding nerve structures in the resected tissue

To gain insight into the appearance of ENS and extrinsic TNBs across the resected tissue, we performed immunofluorescence for p75 which is expressed by both cells of the ENS and perineurial cells of the TNB [6,14], and GLUT1, the TNB perineurial marker. We detected p75-positive cells within the ganglia in the myenteric and submucosal plexus (Figure 1a, b, Supplementary Figure 2) in the most proximal, ‘normoganglionic’ segment (segment I). Negligible GLUT1 staining was found in the enteric ganglia in this segment (Fig. 1b). Progressing distally, in segment II we also detected p75-expressing cells throughout the ganglia. In contrast to the ganglia in segment I, strong and distinct GLUT1 expression was seen in a ring-like structure surrounding the submucosal ganglia (Figure 1c, Supplementary Figure 2); a small number of GLUT1-positive cells were also apparent at the periphery of the myenteric ganglia in this segment (Fig. 1d). In segments III and IV, p75-positive neural structures were bounded by a perineurium which strongly stained for both GLUT1 and p75 (Fig. 1e-h). Higher powered confocal microscopy revealed that the cells co-expressing p75 and GLUT1 intimately surrounded the tightly packed neuronal structures in segments III and IV (Figure 2a, b). Due to their size and distinctive p75 staining these could be identified as TNBs.

**Figure 1.**
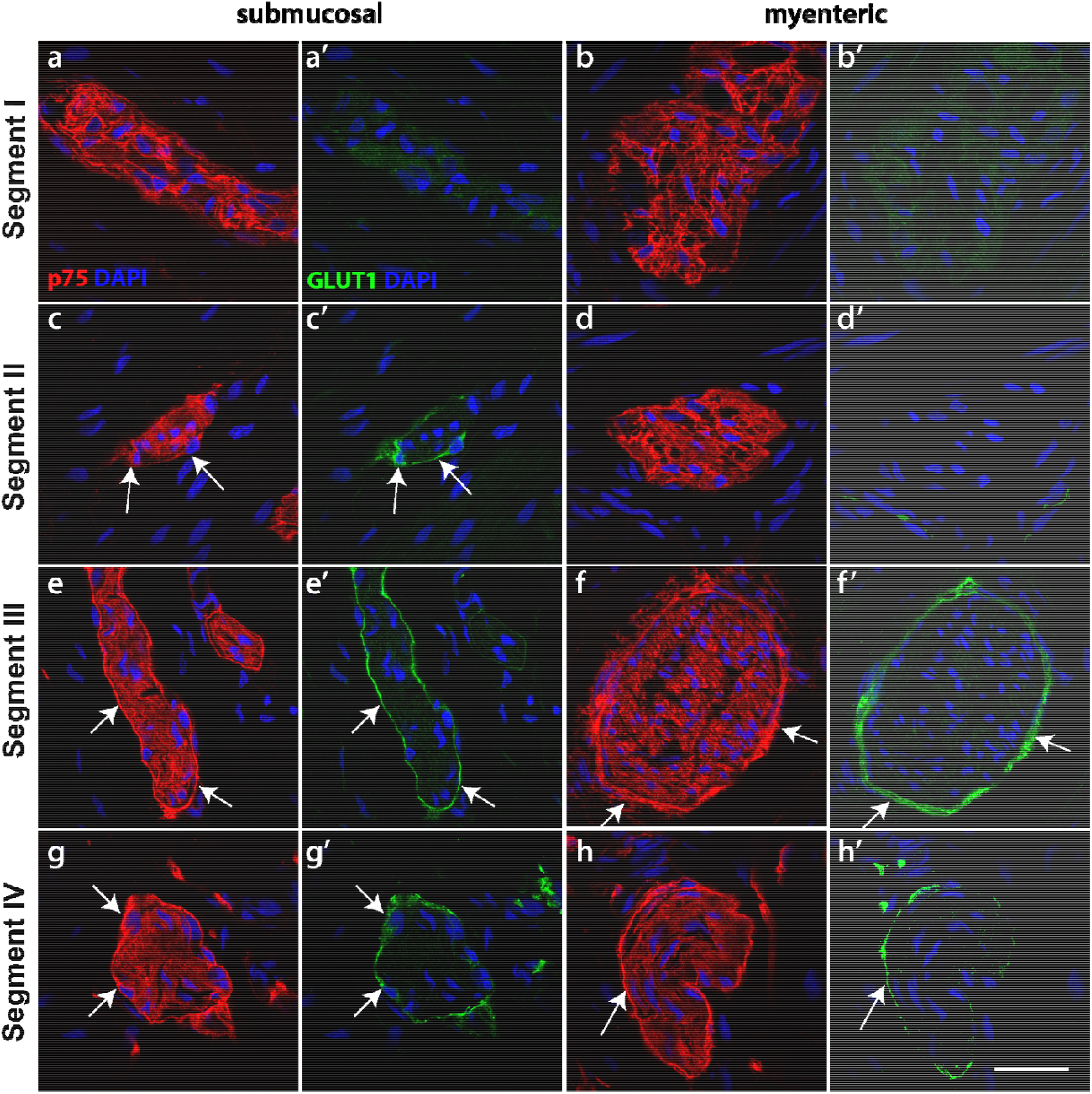
p75 and GLUT1 immunofluorescence through the resected tissue. Tissue sections from HSCR pullthrough colon were stained with p75 (red) and GLUT1 (green). Images show both the submucosal plexus (a,c,e,g) and the myenteric plexus (b,d,f,h). In segment I (a,b), p75 is expressed thoughout the ganglia, but GLUT1 was absent (a’,b’). Enteric ganglia in the myenteric plexus of segment II (d) expressed p75 but not GLUT1, while ganglia in the submucosal plexus of segment II (c) showed p75 expression throughout and also revealed a p75-positive perineurium (arrows). This perinerium was also GLUT1-positive (c’, arrows). In segments III and IV (e-h), nerve bundles displayed both a distinctive p75-positive perineurium (arrows), and p75-positive fibres within the TNB. In each case, the perineurium was also labelled by GLUT1 (e’-h’). Representative images of results from 4 patients. Scale bar = 20μm, for all images.

**Figure 2.**
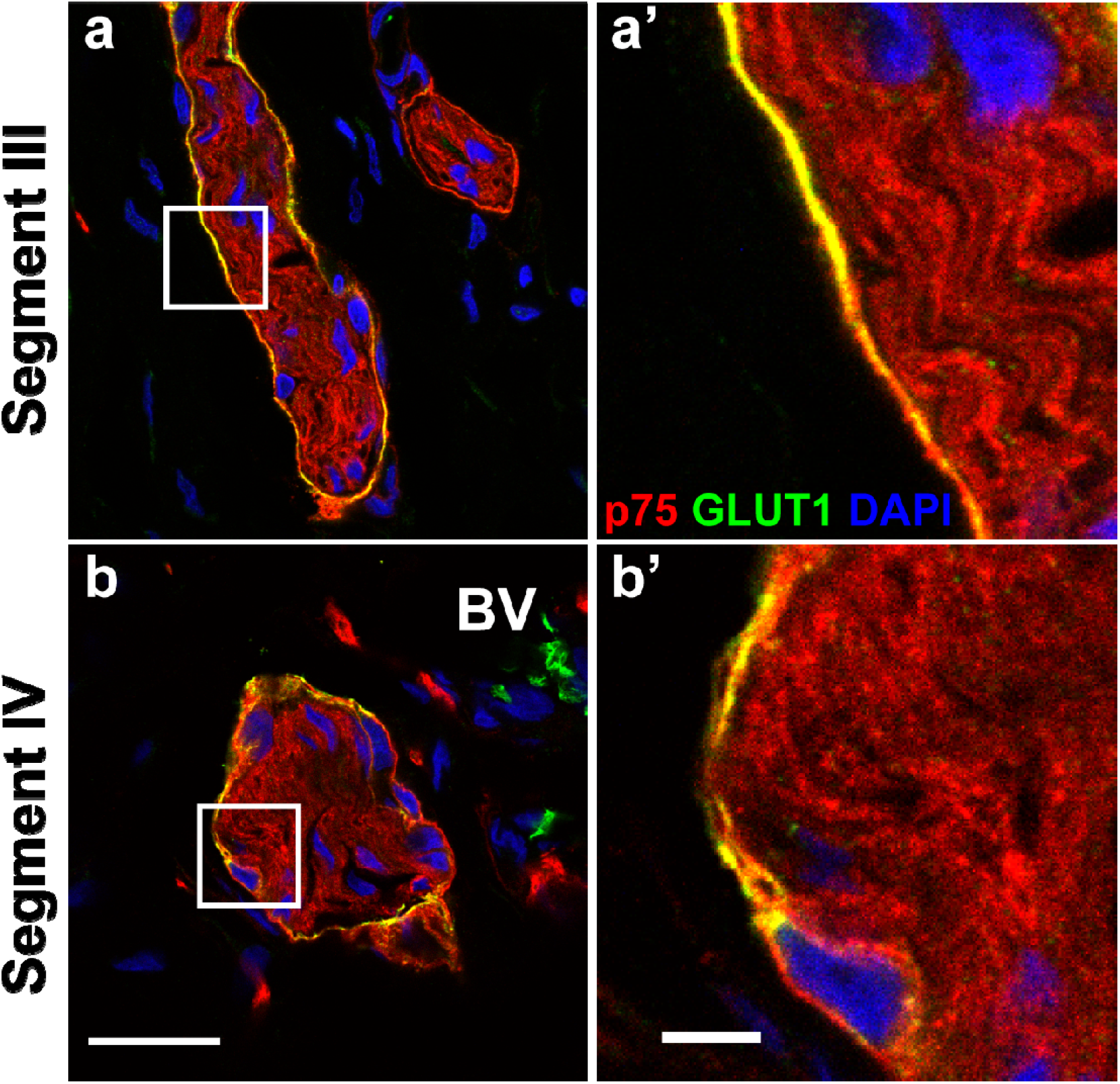
Confocal images of the dual staining of the perinerium with p75 and GLUT1 in Hirschsprung’s bowel. Confocal images and magnified views of the nerve bundles in segment III (a) and segment IV (b) highlight co-expression of p75 (red) and GLUT1 (green) in the perineurium. GLUT1 was not found within the nerve fibres in either segment. Images shown in (a, b) are same as those shown in Figure 1e, g. Scale bar for a, b (as shown in b) = 20μm, scale bar for a’, b’ (as shown in b’) = 10μm.

By combining immunofluorescence staining for the pan-neuronal marker Tuj with p75, we confirmed that the structures in segments III and IV contained nerve fibres and were not enteric ganglia (Supplementary Figure 3), defining segment III as proximal aganglionic bowel. These data indicate that both segment III and IV lacked enteric ganglia and were aganglionic. Our analysis also revealed that the ring-like strong expression of p75 throughout segments II-IV was not observed for Tuj.

These observations demonstrate the spatial context of the changes to the ENS through the resected tissue of SS-HSCR colon by showing that ENS ganglia were lost, and extrinsic nerve bundles appeared which were ensheathed by a perineurium that co-expressed GLUT1 and p75.

### Hu-positive neuronal cell bodies are found in hybrid structures in the transition zone

To determine the extent of the presence of ENS neuronal bodies in the neuronal structures displaying both ENS and TNB markers in segments II and III, we labelled for the neuronal marker Hu in combination with p75 (Figure 3, Supplementary Figure 4). Hu is only expressed in cell bodies and not the fibres or axons of nerves [10,25]. Our data indicate that neuronal structures in both myenteric and submucosal plexus of segment II contained Hu-expressing cell bodies of ENS neurons (Figure 3c, Supplementary Figure 4b). We observed p75 expression in faint perineurial structures only in the submucosal plexus of segment II. By contrast, neuronal structures in segment III lacked Hu staining, indicating the absence of neurons (Figure 3e-h), but were characterised by p75-positive perineurium (Figures 1, 2). These observations suggest that neuronal structures in submucosal segment II contained both perineurial cells and ENS cell bodies, which we consequently define as hybrid structures, and the segment as true transition zone.

**Figure 3.**
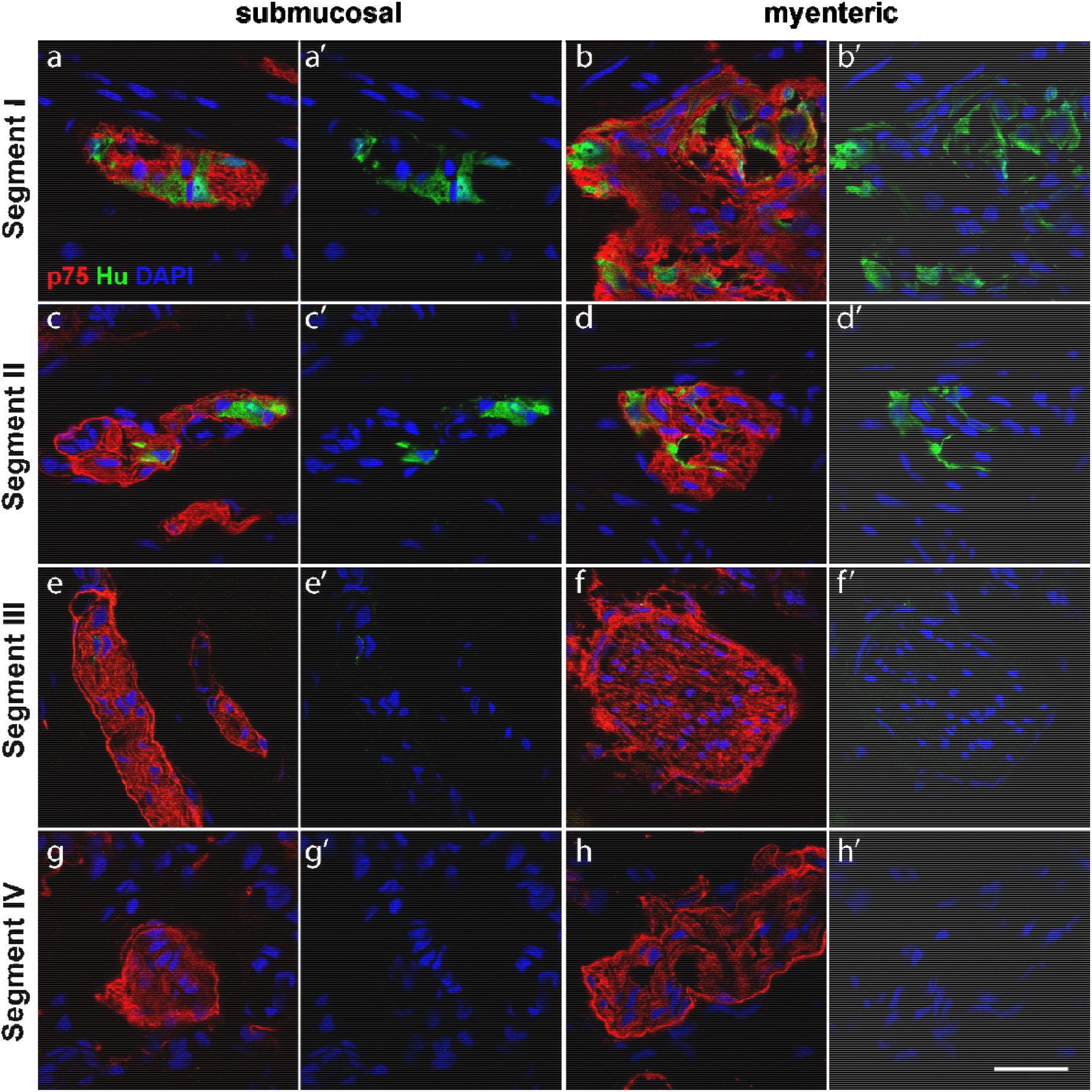
p75 and Hu immunofluorescence through the resected tissue. Tissue sections from HSCR pullthrough colon were stained with p75 (red) and Hu (green). Images show the submucosal plexus (a,c,e,g) and the myenteric plexus (b,d,f,h). In segments I (a,b) and II (c-d), p75- and Hu-expressing cells are present throughout enteric ganglia of both plexus as well as in the submucosal hybrid structure (c) as defined by the p75-positive perineurium. In segments III and IV (e-h), there is an absence of Hu expression. Representative images of results from 4 patients. Scale bar = 20μm, for all images.

### The ENS neuronal marker Calretinin is expressed in hybrid structures in the transition zone

We next asked whether the hybrid structures containing a perineurial sheath and Hu-positive neuronal cell bodies, as observed in segment II, had characteristics of enteric ganglia. We addressed this by labelling for calretinin, a specific marker for a subset of ENS neurons [26] in combination with p75. Our results show that enteric ganglia in segment I contained calretinin-positive neurons and no p75-positive perineurial cells (Figure 4a, b). By contrast, in the submucosal plexus of segment II we found calretinin-expressing cells that were surrounded by p75-positive perineurium (Figure 4c). Of note, the calretinin-expressing cells in the myenteric plexus in segment II lacked a ring of p75-positive perineurium (Figure 4d). We also observed perineurial structures surrounding calretinin-positive enteric neurons in both the submucosal and myenteric plexus of segment III (Figure 4e, f), suggesting their hybrid characteristics. However, in segment IV we failed to detect any calretinin-positive cells within the perineurium-bounded TNBs (Figure 4g, h), demonstrating a gradual loss of calretinin-expressing ENS neurons across the region.

**Figure 4.**
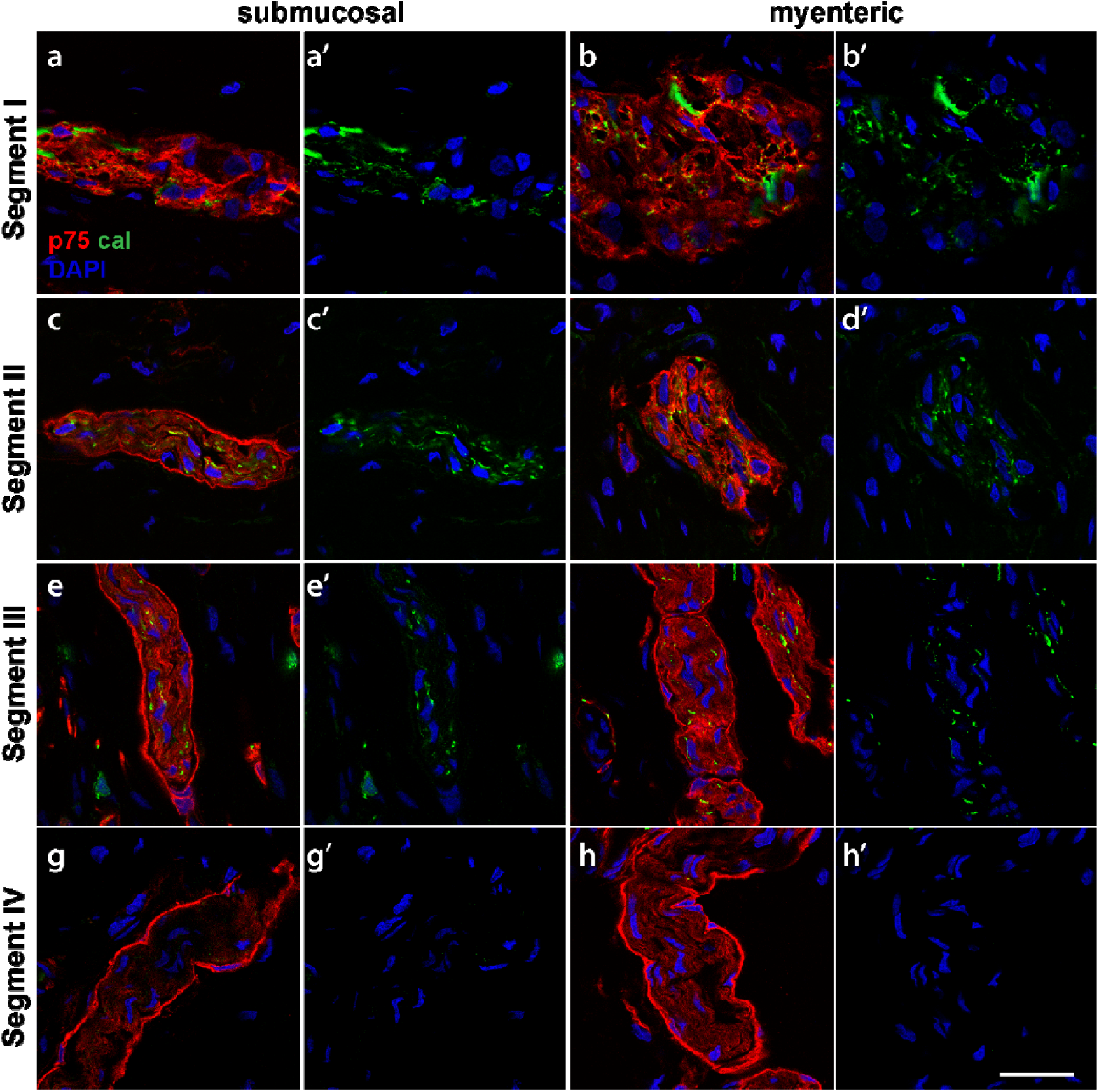
p75 and calretinin immunofluorescence through the resected tissue. Tissue sections from the HSCR colon were stained with p75 (red) and calretinin (green). Images show the submucosal plexus (a,c,e,g) and the myenteric plexus (b,d,f,h). In segment I (a,b), p75 and calretinin were present throughout ganglia of both plexus. In segment IV (g,h), there is an absence of calretinin expression while p75-positive cells were present in the perineurium. Hybrid structures were observed in segments II and III (c-f). In these regions, there were nerve bundle structures containing p75-positive perineurium, with calretinin positivity within. Representative images of results from 4 patients. Scale bar = 20μm, for all images.

Using confocal imaging, we analysed the calretinin distribution across the neuronal structures in segments I-III (Figure 5). While entire cells were positive for calretinin in segment I (Figure 5a, b), we found only punctate expression of calretinin in segments II and III in both myenteric and submucosal plexus (Figure 5c-f).

**Figure 5.**
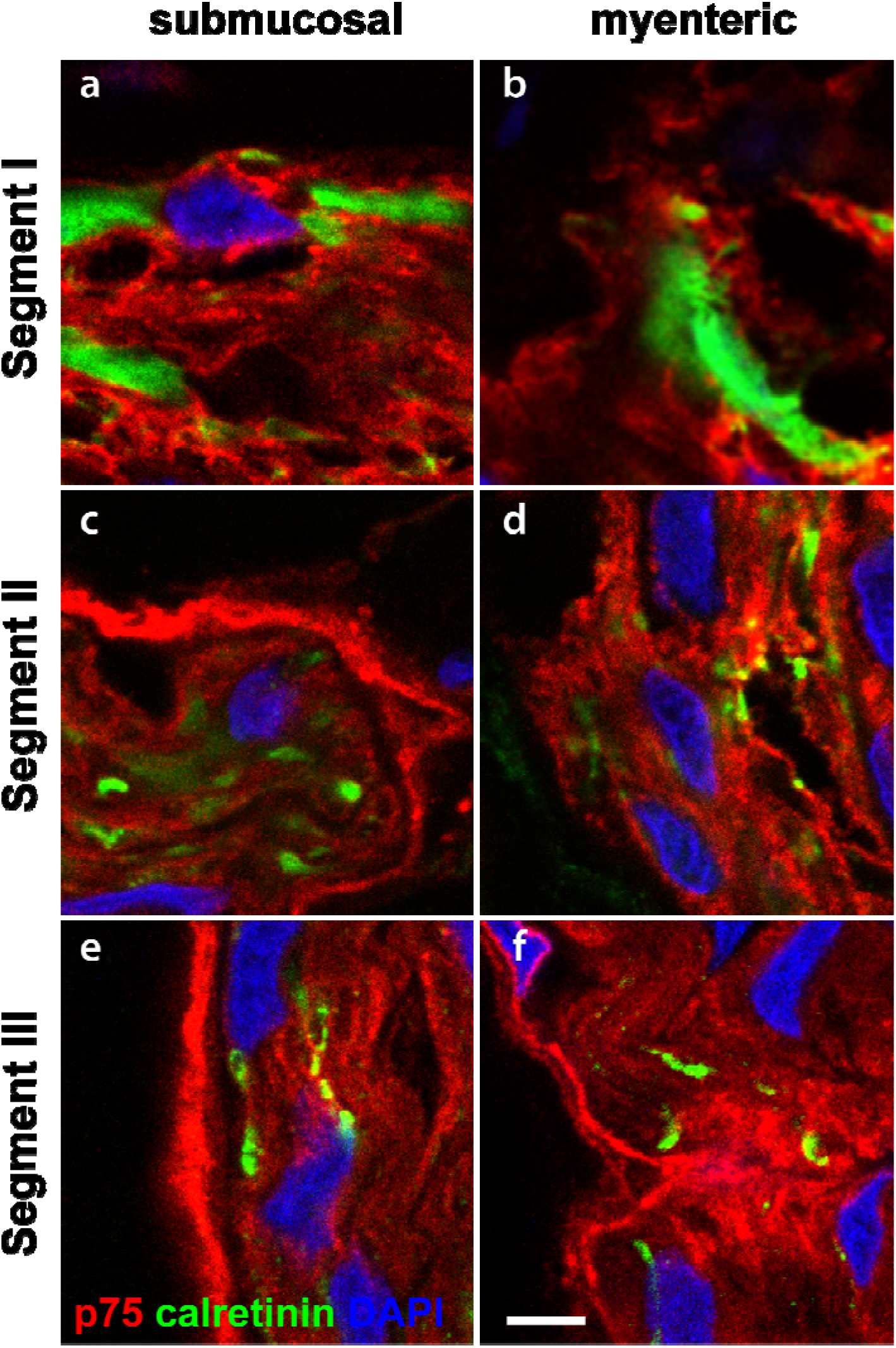
Confocal images of the dual staining of the perinerium with p75 and calretinin in Hirschsprung’s bowel. Magnified views of confocal images of the enteric ganglia and hybrid structures found in segment I, segment II and segment III with p75 (red) and calretinin (green). Calretinin was found in cell bodies in segment I, but in the more distal segments became restricted to a more lineate punctate pattern, suggesting its presence in cell projections. Scale bar = 10μm, for all images.

It is important to note that calretinin is expressed in both the cell bodies and axons of a subset of enteric neurons [26,27]. This suggests that calretinin-positive staining in hybrid structures in segment III (Figure 4, 5) may have detected the axons of ENS neurons that extended caudally within extrinsic nerve bundles bounded by a perineurial sheath. Our results also suggest that there was a difference in the cranio-caudal change of ENS ganglia to hybrid structures between myenteric and submucosal plexus in the TZ: hybrid structures appeared in segment II in the submucosal plexus while in the same segment the myenteric plexus still displayed typical ENS ganglia lacking a perineurial sheath.

### Localization of telocytes in enteric ganglia, hybrid structures and TNB in the resected Hirschsprung’s bowel

To determine the extent to which the hybrid structures in the TZ have a normal telocyte ensheathment despite the presence of an abnormal perineurium, we co-labelled sections across segments I-IV with antibodies for the telocyte marker CD34 (Figure 6, Suppl Figure 5). Telocytes were found surrounding normal enteric ganglia in segment I (and II), as previously reported [17], in a continuous, uninterrupted sheath, extending telopode processes into the interior of the ganglia (Figure 6a, b, Supplementary Figure 5). By contrast, telocytes surrounded the hybrid structures in segment III and TNBs in segment IV in a discontinuous layer on both the inner side of the p75-positive perineurium and outside of the perineurium (Figure 6c-d, Figure 7). This pattern reflected that of extrinsic nerves as previously reported [21].

**Figure 3.**
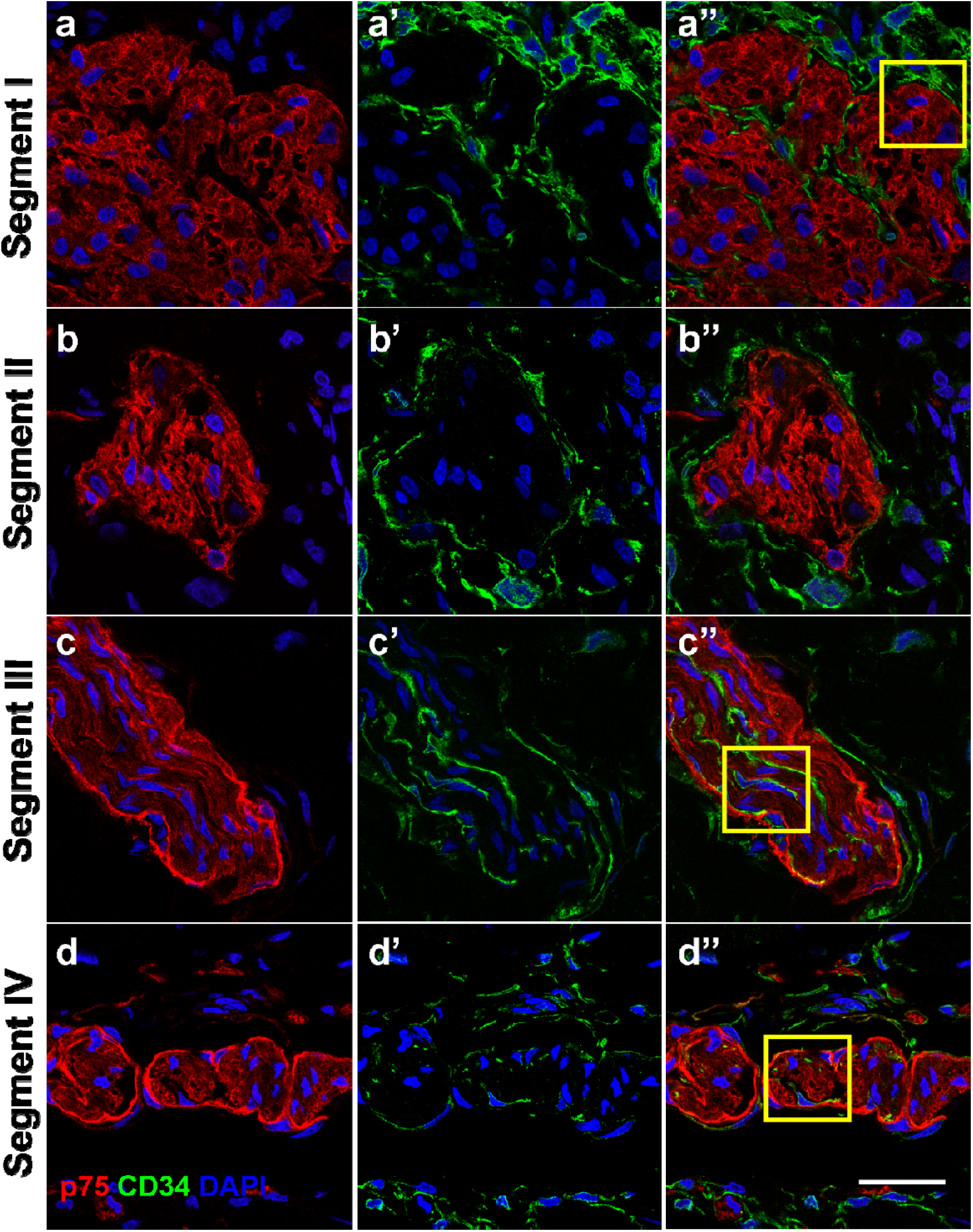
p75 and CD34 immunofluorescence staining in the myenteric plexus of Hirschsprung’s bowel. Tissue sections from HSCR colon were stained with p75 (red) and CD34 (green) in the myenteric plexus. In segments I and II, CD34 can be found to make a complete layer around the enteric ganglia. Telopodes also extent into the ganglia in segment I. In segments III and IV, CD34 positive cells are found outside within the p75 positive perineurium, as well as along the nerve fibres. Telocytes are also found throughout the muscle layers. Representative images of results from 4 patients. Yellow boxes indicate field of view shown in Figure 7. Scale bar = 20μm, for all images.

**Figure 4.**
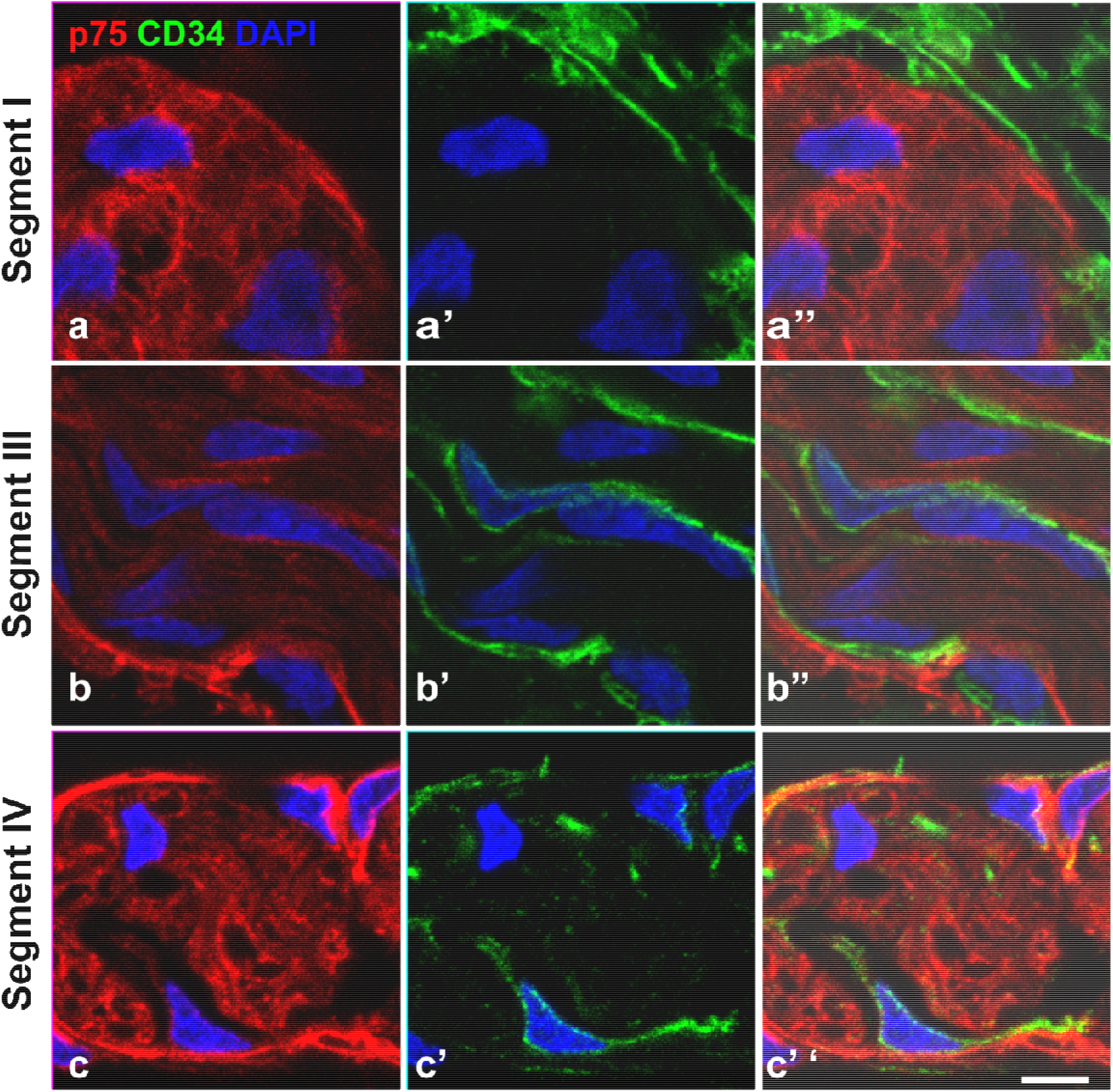
Confocal images of the dual staining of the perineurium with p75 and CD34 in Hirschsprung’s bowel. Magnified views of confocal images of the ganglia and hybrid structures and TNBs found in segment I, segment III and segment IV with staining for p75 (red) and CD34 (green). In segment I, a close contact between the ganglion and telocytes is shown. In segment III, and IV, telocytes were found close to the perineurium of the nerve bundles and along the nerve fibres. Representative images of results from 4 patients. Images are magnified fields of view as shown in Figure 6 a’’,b’’,c’’. Scale bar = 10μm, for all images.

## Discussion

In order to elucidate cellular relationships between the TNB and ENS, we have focussed on the transition zone between ganglionic and aganglionic regions of the HSCR bowel because previous observations have demonstrated that enteric ganglia and TNB co-exist within the TZ [10,23]. However, any direct cellular contact between enteric ganglia and extrinsic nerves has not previously been shown.

Our analysis of four cranio-caudal segments along the transition zone of HSCR bowel confirms that the ENS and TNB undergo gradual reciprocal changes from ganglionic to aganglionic zones [10,23]. This gradual loss of the typical ganglia is accompanied by the appearance of hybrid structures which contain cells expressing ENS markers and at the same time are surrounded by GLUT1-positive perineurium characteristic of TNB and other extrinsic nerves. However, it remains to be determined if perineurium of the abnormal ganglionic hybrid structures is derived from the TNB, or alternatively, if the environment within the postnatal TZ or in the gut during prenatal development induces peripheral cells of ENS ganglia to adopt a perineurial phenotype.

While 3 of the 4 patients from which tissue was collected underwent pull-through without prior surgery, one child had previously had a stoma placed before the pull-through surgery from which the tissue was collected. It has been demonstrated in neonatal rats [28] that transection of the bowel to form a colostomy can change the morphology of the ENS in the distal colon. Govaert et al., [28] demonstrated that the transection of the bowel caused a change in the composition of neurons and ganglia of the ENS due to a lack of activity in the bowel. Despite these findings in a rat model, there has been no subsequent evidence for a similar phenomenon in humans. Kapur and colleagues investigated if there was remodelling of the bowel following pull-through surgery in HSCR patients, specifically if extrinsic nerves could innervate the neorectum but found no such remodelling of the neorectum from native pelvic nerves [29]. Given that the results of the one patient with a previously placed ostomy did not differ from the other patients, we do not think that this difference affected the morphology/ innervation of the TZ.

During development, the perineurium of peripheral nerves is formed from specialised Schwann cells [30,31] that surround and ensheath extrinsic nerve fibres in the gut. The perineurium is formed in two main stages: in the first step, mesenchymal cells generate a loose permeable sheath; in a second step, this primitive sheath develops to form a multi-layered structure via desert hedgehog (Dhh) signalling from Schwann cells, present inside the nerve fibres, that then contribute to the perineurium itself [30,31]. Both GLUT1 and p75 have been shown to be strongly expressed in the perineurium [10,26], while only occasional weak staining of GLUT1 has been reported within enteric ganglia [6,26]. While extrinsic nerve fibres have been reported in SS-HSCR biopsies based on the GLUT1 and p75 expression in the perineurium [10], an analysis of their distribution and arrangement in the transition zone, has not previously been reported. Our analysis shows that a GLUT1- and p75-expressing perineurium appears as a discrete boundary around the ENS neurons of hybrid structures and is detectable in the submucosal plexus at more cranial levels closer to the normoganglionic zone than in the myenteric plexus where they are restricted to more caudal regions closer to the aganglionic gut in HSCR patients.

In order to understand the relevance of the hybrid structures it is important to revisit the developmental process in which the ENS precursor cells migrate, populate and differentiate within the bowel. In the mouse, it has been shown that vagal neural crest cells colonise the myenteric plexus first and begin to differentiate into neurons before radial migration and projection of ENS precursor cells and Schwann cell precursor cells into the submucosal plexus [32-34]. It has been suggested that in HSCR the development of the ENS is arrested at the stage where migration of vagal neural crest cells and differentiation of ENS precursor cells from the myenteric plexus towards the submucosal is halted [35]. Therefore, the appearance of hybrid structures in the submucosal plexus more cranially when compared to the myenteric, may suggest that the development of the enteric ganglia in the submucosal plexus of had not progressed [32,36,37]. Instead, these incomplete enteric ganglia contribute to the formation of hybrid structures, which may represent the point at which extrinsic nerves have reached the submucosal plexus.

A hybrid-like structure has been previously described in a colon biopsy by Kakita and colleagues [6], where ganglion cell bodies were surrounded by a perineurium. The authors suggested that this observation could serve as a histopathological diagnostic feature of patients with intestinal neuronal dysplasia (IND) rather than HSCR [6]. IND is a poorly studied condition [38] which has been reported to present with features similar to HSCR, such as intestinal obstruction and histological structures consisting of ganglion cell bodies with a perineurium [38,39]. There has in fact been doubt as to the existence and/ or clinical significance of IND due to there being a lack of consistency in diagnostic criteria, and it therefore remains a controversial diagnosis [40]. It needs to be determined whether the hybrid-like structure described by Kakita as IND [6], has a similar developmental origin to hybrid structures in the transition zone of HSCR that we describe here.

A detailed analysis of the four segments of HSCR pull-through resected colon tissues using the neuronal markers calretinin, Hu and Tuj provided insight into the extent of intrinsic innervation across these tissues. Staining with calretinin visualised the change from cytoplasmic expression in the ENS neurons in the ganglionic bowel in segment I to a more granular localisation more distally (Segments II and III) before its absence in the aganglionic bowel in segment IV. While calretinin is only found in a subset of enteric neurons, nevertheless these data indicate, as previously hypothesised [41], that calretinin-expressing neuron cell bodies identified by Hu staining project axons caudally from the rostral segments I and II into the proximal aganglionic segment III.

It has been shown that in mouse models of HSCR, neurogenesis can occur from Schwann cell precursors (SCPs) [42], a source of enteric neuron progenitors [43]. SCP-derived neurons appear to be most abundant in the TZ [42], where there is a reduction in the presence of vagal neural crest cells. Therefore, the calretinin- and Hu-positive cells that we have detected in the TZ could be interpreted as SCP-derived neurons.

Here, we have shown that telocytes ensheath the enteric ganglia in segments I and II, in line with their reported arrangement in normal enteric ganglia [17]. We further demonstrate that telocytes persist in the aganglionic region of HSCR gut where they align with and integrate within the perineurium of TNBs, consistent with reports that telocytes are present in the endoneurium as well as either side of the epi- and perineurium of extrinsic nerves [21]. Our observations show that telocytes are present in the caudal segments of resected aganglionic bowel where they are surrounded by the perineurium-bounded TNB, similar to the arrangement in extrinsic nerves. Additionally, the telocyte distribution in segments II and III demonstrates their presence within the hybrid structures that have both ganglia-like and extrinsic nerve-like properties. Specifically, telocytes form a near continuous layer around hybrid structures, similar to ganglia, but are also found within the perineurium and within the structure similar to that of TNBs and as described in peripheral nerves [21]. How telocytes undergo the changes in the hybrid structures in the TZ of SS-HSCR bowel remains unclear. Further studies are required to dissect how the presence and function of telocytes in HSCR bowel is regulated.

Telocytes have a range of roles including the regulation of GI motility along with ICCs [16] by forming networks that provide mechanical support for connective tissue components in peristalsis and maintain integrity of ganglia by entirely ensheathing them. In this role, telocytes are thought to be involved in the 3D-organisation of the extracellular matrix that embeds the ENS. Interestingly, in Crohn’s disease and ulcerative colitis the numbers of telocytes are reduced [17,44,45], which is accompanied by the excessive production of a disorganised extracellular matrix, leading to fibrotic remodelling of the bowel wall [19] and a discontiunous network of telocytes around enteric ganglia [45], as seen in the TZ and aganglionic bowel. It has been suggested that the disorganisation of telocytes contributes to the colonic dysmotility and derangement of intestinal wall observed in Crohn’s disease and Ulcerative Colitis [44,45], which may also be the case in HSCR. Here, we have observed that the continuous arrangement of the telocytes which surround normal ENS ganglia is disrupted and discontinuous in hybrid structures and extrinsic TNBs. While the arrangement of telocytes in TNBs is comparable to that of peripheral nerves, the loss of a complete sheath of telocytes, as is present around ganglia in normoganglionic tissue, could impact the organisation and arrangement of the extracellular matrix, or contribute to colonic dysmotility along with the lack of ENS in aganglionic bowel.

Further analyses are required to determine the aetiology of the hybrid structures in the transition zone of HSCR, and the role of telocytes in ENS ganglia and in the perineurium of TNBs.

## Data Availability

All data produced in the present study are available upon reasonable request to the authors

## Acknowledgements

We acknowledge funding for a PhD studentship for MS from the Bowel and Cancer Research Charity (now Bowel Research UK) and the Institute of Integrative, Systems and Molecular Biology (University of Liverpool), for a project grant from the Children’s Research Fund for MS, as well as a Medical Research Council-funded clinical research training fellowship MR/R002/42/1 for SC. We acknowledge use of the Centre for Cell Imaging facilities provided by the Liverpool shared Research Facilities, Faculty of Health and Life Sciences, University of Liverpool.

## Author Contribution statement

MS, SC, BW, DE designed experiments. Tissue collection, immunofluorescence staining and imaging were carried out by MS and SC. Figures were prepared by MS. Manuscript was written by MS, BW, DE and all authors approved manuscript before submission.

## Data statement

Data will be made available upon request.

