## Supplementary materials for "The transition zone in Hirschsprung’s bowel contains abnormal hybrid ganglia with characteristics of extrinsic nerves"

**Supplementary figures**

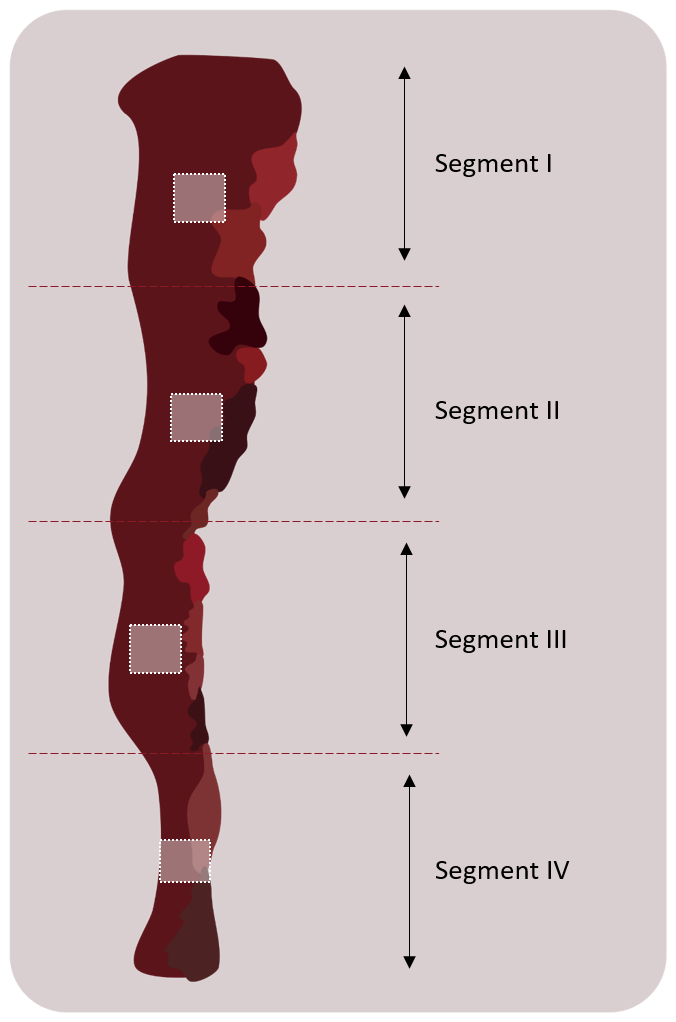

**Supplementary Figure 1. Schematic illustrating the tissue resected from pull-through surgery.** From the tissue resected, 4 pieces of tissue were collected. A small piece of tissue each was taken from 4 regions, labelled segment I-IV.

**Supplementary Table 1**: Primary antibodies for immunofluorescence staining

| Antibody | Description | Location | Host/Details | Dilution | Supplier |
| --- | --- | --- | --- | --- | --- |
| Anti-p75 NGF Receptor (NGFR5) | Enteric neural crest progenitor marker | Cell surface | Mouse IgG1  Monoclonal | 1:500 | Abcam (ab3125) |
| Anti-Calretinin | Enteric Neuronal marker (subset of enteric neurons) | Cytoplasm (ganglia) | Rabbit IgG1 Monoclonal | 1:50 | Abcam (ab702) |
| Anti-HuD + HuC | Human neuronal marker | Cell soma/ cytoplasm | Rabbit Monoclonal | 1:500 | Abcam (ab184267) |
| Anti-glucose transporter 1 (GLUT1) | Expressed in nerve perineurium | Cell membrane | Rabbit IgG polyclonal | 1:100 | Abcam (ab15309) |
| Anti-CD34 | Telocyte marker | Membrane | Recombinant Rabbit monoclonal | 1:250 | Abcam  (ab81289) |
| Anti-Tubulin β3 (Tuj) | Neuronal microtubule marker | Cytoskeleton | Mouse IgG2a monoclonal | 1:500 | Biolegend (801201) |

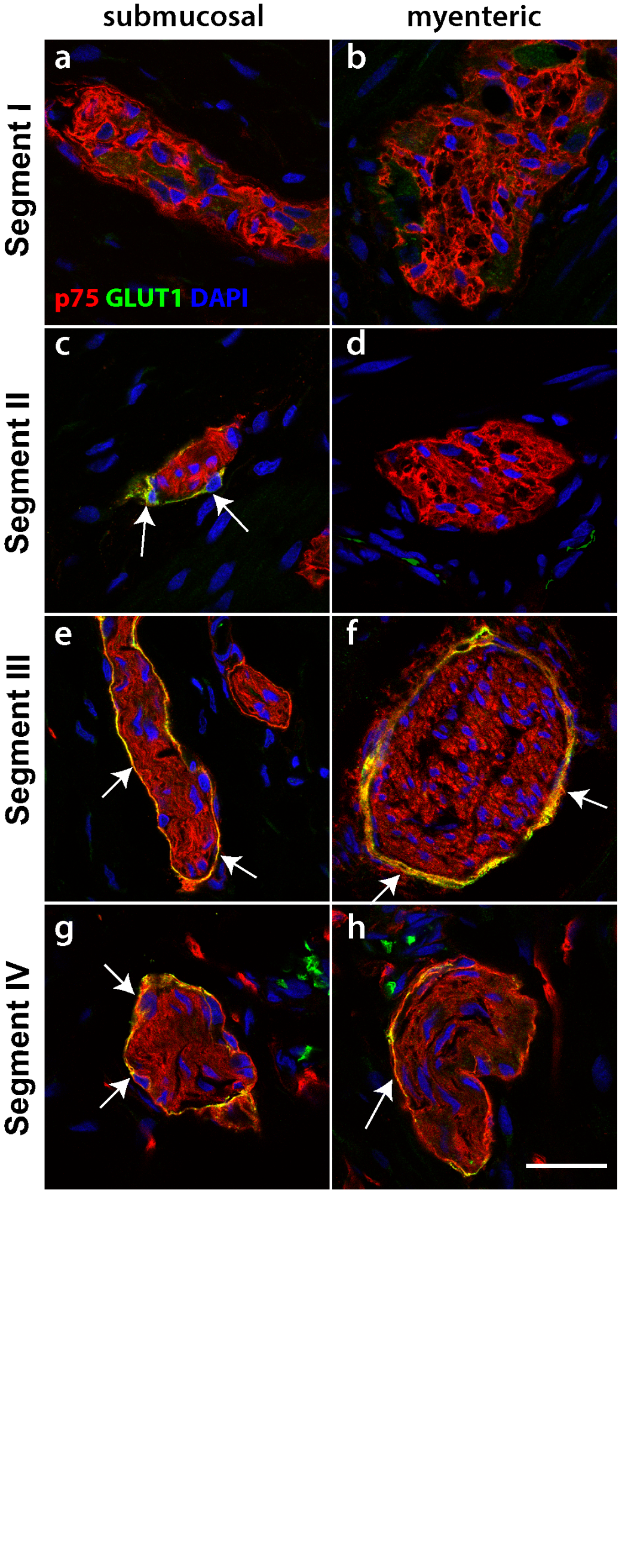

**Supplementary Figure 2. p75 and GLUT1 immunofluorescence through the resected tissue.** Tissue sections from HSCR pullthrough colon were stained with p75 (red) and GLUT1 (green). Images show both the submucosal plexus (a,c,e,g) and the myenteric plexus (b,d,f,h). In each case, the perineurium was also labelled by GLUT1 (e-h). Representative images of results from 4 patients. Scale bar = 20µm, for all images.

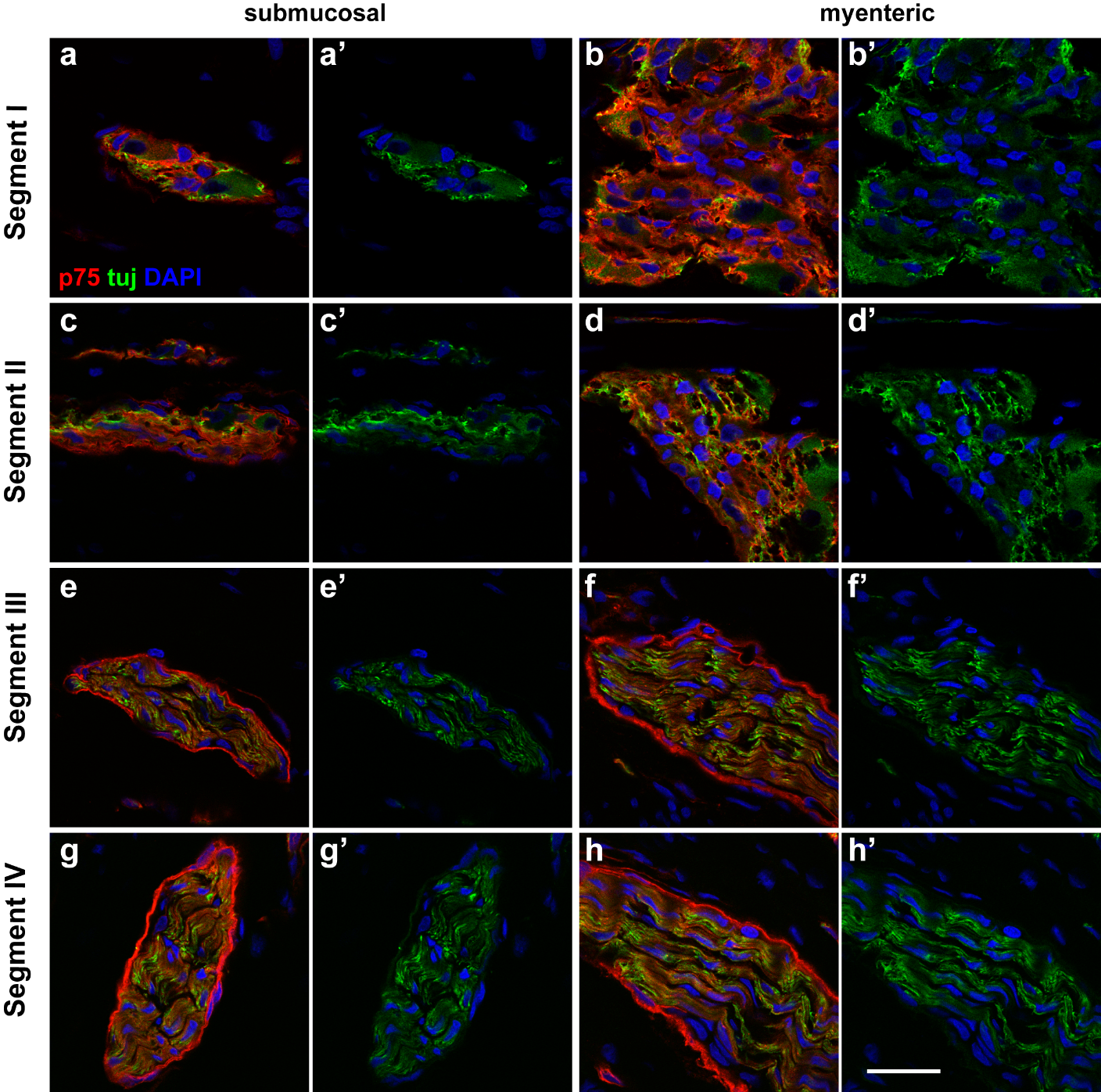

**Supplementary Figure 3. p75 and Tuj immunofluorescence staining through the resected tissue.** Tissue sections from the HSCR colon were stained with p75 (red) and Tuj (green). Images show the submucosal plexus (a,c,e,g) and the myenteric plexus (b,d,f,h). In segments I (a,b) and II (c,d), p75 and Tuj were present throughout ganglia of both plexus. In segments III (e,f) and IV (g,h), while the perineurium was only stained with p75 and not Tuj, the Tuj staining was present in a nerve fibre pattern as opposed to the staining throughout the ganglia. Representative images of results from 4 patients. Scale bars = 20µm for all images.

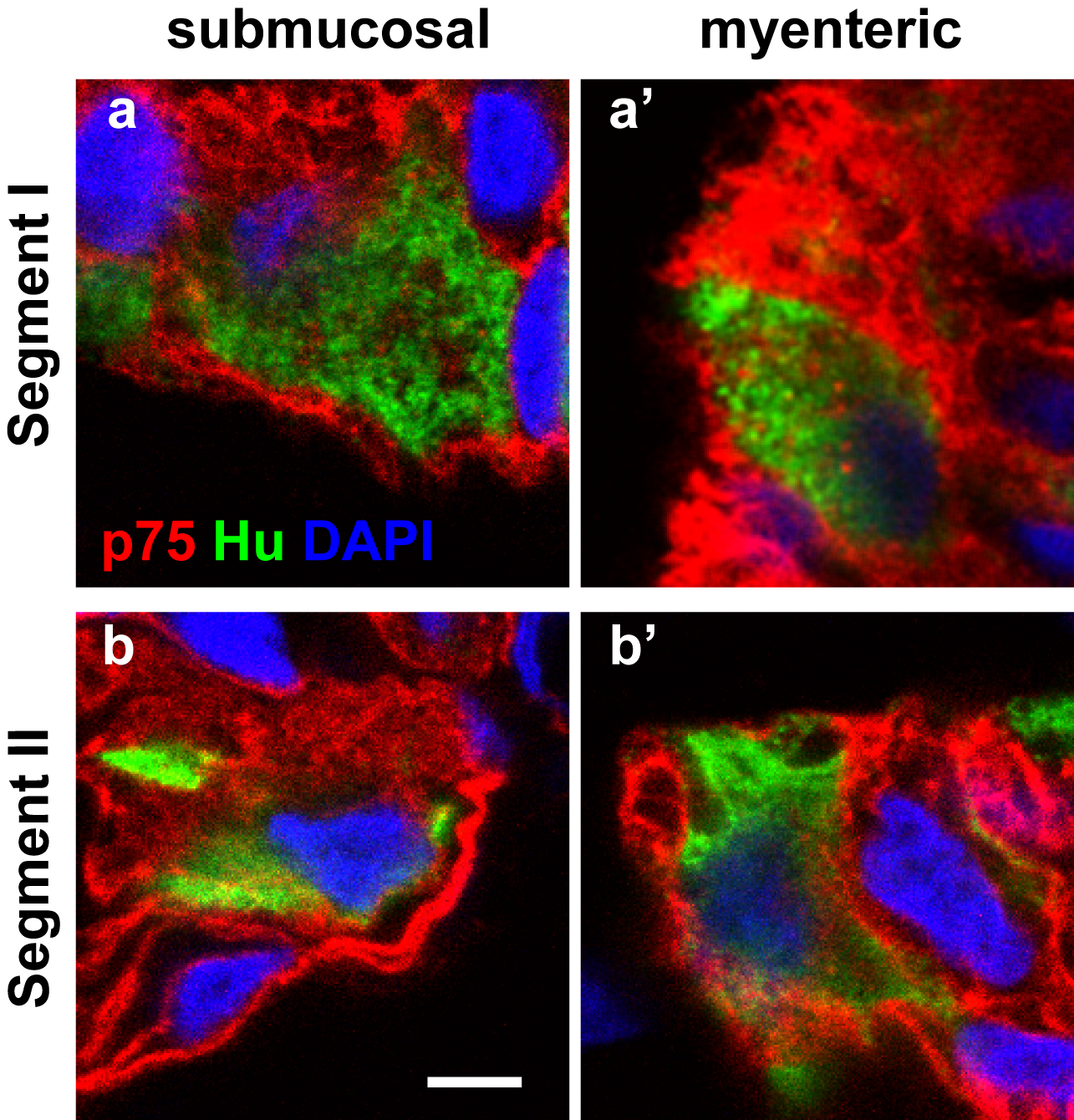

**Supplementary Figure 4. Magnified images of the dual staining of ganglia and hybrid with p75 and Hu in Hirschsprung’s bowel.** Magnified views of confocal images of the ganglia and hybrid structure found in segments I and II labelled for p75 (red) and Hu (green). Hu was found in cell bodies in both ganglia (a, a’, b’) and within the hybrid structure (b). Scale bar = 20µm for all images.

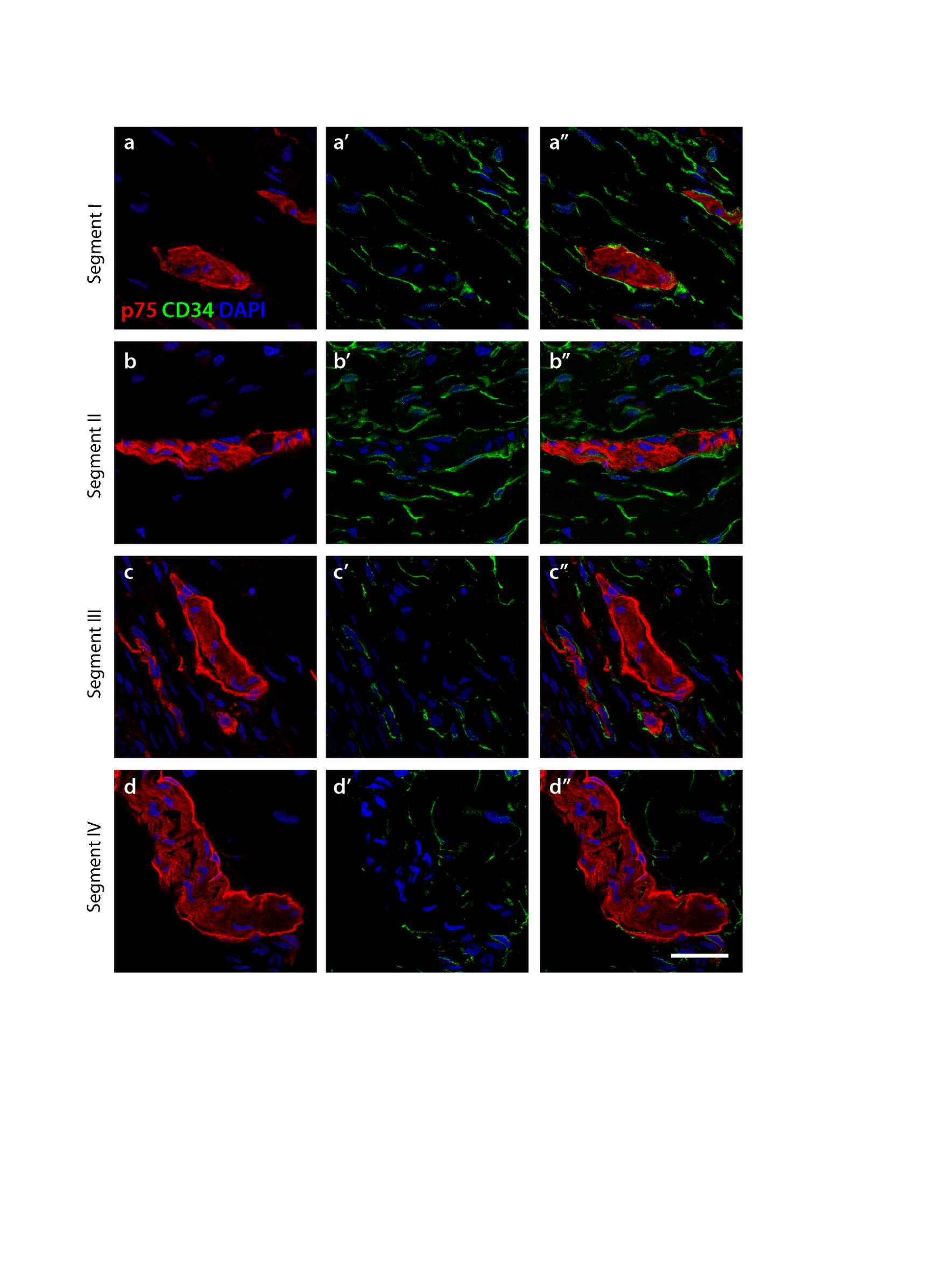

**Supplementary Figure 5. p75 and CD34 immunofluorescence staining in the submucosal plexus of Hirschsprung’s bowel.** Tissue sections from HSCR colon were stained with p75 (red) and CD34 (green) in the submucosal plexus. In segment I, CD34-positive telocytes can be found to form a complete layer around the ganglia, and telopodes extend into the ganglia. In segment II, the telocytes layer is distrupted. In segments III and IV, CD34-positive cells are found outside the p75 positive perineurium, as well as along the nerve fibres. Representative images of results from 4 patients. Scale bars = 20µm for all images.
